# Multi-strain carriage and intrahost diversity of *Staphylococcus aureus* among Indigenous adults in the US

**DOI:** 10.1101/2024.06.07.24308612

**Authors:** Julia Webb, Eleonora Cella, Catherine Sutcliffe, Catherine Johnston, Sayf Al-Deen Hassouneh, Mohammad Jubair, Dennie Parker Riley, Carol Tso, Robert C. Weatherholtz, Laura L. Hammitt, Taj Azarian

## Abstract

*Staphylococcus aureus* (SA) is an opportunistic pathogen and human commensal that is frequently present in the upper respiratory tract, gastrointestinal tract, and on the skin. While SA can cause disease ranging from minor skin infections to life-threatening bacteremia, it can also be carried asymptomatically. Indigenous individuals in the Southwest United States experience high rates of invasive SA disease. As carriage is the most significant risk factor for disease, understanding the dynamics of SA carriage, and in particular co-carriage of multiple strains, is important to develop strategies to prevent transmission in vulnerable communities. Here we investigated SA co-carriage and intrahost evolution by performing high density sampling from multiple anatomical sites and whole-genome sequencing (WGS) on 310 SA isolates collected from 60 Indigenous adults participating in a cross-sectional carriage study. We assessed the richness and diversity of SA isolates via differences in multi-locus sequence type (MLST), core-genome single-nucleotide-polymorphisms (SNPs), and genome content. Using WGS data, we identified 95 distinct SA populations among 60 participants; co-carriage was detected in 42% (25/60). Notably, two participants each carried four distinct SA strains. Variation in antibiotic resistance determinants among carried strains was identified among 42% (25/60) of participants. Lastly, we found unequal distribution of clonal complex by body site, suggesting that certain lineages may be adapted to specific anatomical sites. Together, these findings suggest that co-carriage may occur more frequently than previously appreciated and further our understanding of SA intrahost diversity during carriage, which has implications for surveillance activities and epidemiological investigations.

## INTRODUCTION

*Staphylococcus aureus* (SA) is a gram-positive opportunistic pathogen frequently carried asymptomatically in the gastrointestinal tract, upper respiratory system, and skin. While SA can cause a variety of invasive and non-invasive diseases, it is asymptomatically carried by 20-30% of the general population [1]. The most common site of carriage is the anterior nares, but there is an increasing focus on the oropharynx as an additional carriage site [2–6]. Carriage is thought to occur in three main patterns over time: persistent, intermittent or occasional, and non-carriage [7]. Persistent carriers generally carry a single strain and have higher carriage density and greater risk of infection. Intermittent/occasional carriers may carry multiple strains at different time points or even concurrently [7].

SA carriage is one of the most significant risk factors for disease [8,9], and as such, understanding the dynamics of carriage is central to investigating transmission and developing effective prevention strategies. While carriage studies are abundant, those that employ sampling of multiple anatomical sites or perform molecular or genomic analysis of more than a single isolate from a participant are far fewer [10–12]. Therefore, instances of co-carriage are likely to be underestimated. Further, while typing methods including pulsed-field gel electrophoresis (PFGE), multi-locus sequence typing (MLST), and *spa*-typing have historically been used to define the population structure of SA and to infer the genetic relatedness among strains [13,14], the increasing application of microbial genome sequencing, has revealed their limited resolution. Through application of pathogen genome sequencing, we now appreciate the breadth of intrahost microbial diversity that can exist during a single episode of carriage or disease [11,15,16]. Yet, due to varying methodologies and definitions of a strain, the frequency of co-carriage remains unclear with prevalence estimates ranging from 9% to 79% (average 34%) [17–20]. The scarcity and vast range of available co-carriage estimates highlight the need for further studies and standardization of a co-carriage definition.

We recently completed a SA carriage study among Indigenous children and adults in the Southwest US, where the rates of invasive disease are significantly higher than the general US population [21,22]. Using sampling of multiple upper respiratory anatomical sites and sequencing of a single SA isolate from each participant, the carriage prevalence of SA and methicillin-resistant SA (MRSA) were 20.7% and 1.7% among children <5 years, 30.2% and 2.8% among adults 18-64 years, and 16.7% and 3.3% among adults ≥65 years, respectively [23]. Here, we expand on this work by sequencing multiple isolates from each anatomical site for each participant to determine the prevalence of co-carriage, characterize intrahost pathogen genetic diversity, and explore the association between carried strains, anatomical site, and strain-strain interactions.

## METHODS

### Study Population and Sample Collection

Sample collection and study population were described in detail by Cella et al. [24]. Briefly, the cross-sectional study included 288 Indigenous children (<5 years) and adults (≥18 years) living in the Navajo Nation and White Mountain Apache Tribal lands. Participants were recruited in 2017 via convenience sampling at well/routine healthcare visits and community events. Swab samples were collected from the anterior nares (AN) and nasopharynx (NP) of child participants, and from the AN, NP, and oropharynx (OP) of adult participants. Isolation of SA from each swab was performed individually using BBL CHROMagar Staph aureus selective media plates (Becton Dickinson, Franklin Lakes, NJ). From positive plates, up to 6 SA colonies (henceforth “isolates”) were plated on a 6-zone blood agar plate. Each isolate was confirmed as SA using BBL Staphyloslide latex agglutination test (Becton Dickinson, Franklin Lakes, NJ), then inoculated into 1 mL Bacto tryptic soy broth (TSB) (Becton Dickinson, Franklin Lakes, NJ) + 20% glycerol mix and stored at -80C.

In the primary study, 91 participants were found to be SA carriers, including 25 children <5 years of age. As the primary focus of this study was to assess intrahost SA diversity across multiple anatomical sites, we focused this analysis on adults (n=66) with samples collected from all three sites. Six adult participants had an insufficient number of sequenced isolates (< 3) for meaningful analysis and were excluded; 60 participants were included in the final analysis.

### Bacterial gDNA Isolation and whole genome sequencing (WGS)

Bacterial DNA extraction and WGS was carried out as previously described [24]. Briefly, isolates were grown overnight in TSB then treated with 0.1 mg/mL lysostaphin. Bacterial genomic DNA was extracted using the Qiagen DNeasy Blood & Tissue kit with 50 mg/mL lysozyme added to the lysis buffer. DNA quality and quantity were assessed using a Qubit 4 Fluorometer. Short-read sequencing libraries were constructed using the Nextera Flex and Illumina DNA Prep kits and sequenced using an Illumina MiSeq with 600-cycle v3 and 500-cycle v2 flow cells. Previously, one isolate per participant also underwent long-read sequencing using Oxoford Nanopore Technologies (ONT) MinION, allowing for closure of most genomes. These high-quality draft assemblies also were included in the present study.

### Bioinformatics and phylogenetics

*De novo* assembly of sequencing reads was performed using Unicycler v0.5.0 [25], and assemblies were annotated using Prokka v1.14.5 [26]. The MLST of each isolate was determined using the tool MLST (https://github.com/tseemann/mlst) and the PubMLST database [27].

Antibiotic resistance and virulence factor presence was assessed using Abricate (https://github.com/tseemann/abricate) with databases ARG-ANNOT [28] and VFDB [29]. *De novo* assemblies were then used for pangenome analysis, which was carried out using Roary v3.13.0 [30]. SNP-sites v2.5.1 [31] was then used to extract polymorphic sites from the core-SNP alignment. IQTree v1.6.12 [32] with the TVM+F+ASC+R2 substitution model [33,34] was used to create a maximum-likelihood (ML) phylogeny. The phylogenetic tree was visualized and annotated using ggTree [35–37] run in RStudio v.4.1 [38]. Statistical analysis of CC and MLST prevalence was performed using a Monte Carlo estimation of Log-Odds Ratio (https://github.com/sayfaldeen/BioinformaticsScripts/blob/main/MC-LOR-comp.py).

To assess intrahost diversity, an initial intrahost reference-based assembly was generated by mapping each participant’s short-read sequenced isolate to the high-quality hybrid-assembled genome using Snippy v4.6.0 (https://github.com/tseemann/snippy). Pairwise SNP-distances were calculated using snp-dists v0.8.2 for each intrahost population comprised of isolates belonging to the same MLST. We then examined the distribution of SNP-distances for each intrahost population, considering the anatomic site of collection and spatial distribution of SNPs in the genome. In instances of co-carriage of distinct strains, for which we had not previously generated a closed genome through hybrid assembly, the closest matching publicly available reference genome was identified using the KmerFinder online tool (https://cge.food.dtu.dk/services/KmerFinder/). Once intrahost populations were defined, we plotted all pairwise SNP distances as well as the mean intrahost SNP distances for each population.

### Strain definitions

Distinct intrahost populations were delineated using MLST and genetic distance measured from the core genome. Strains were defined initially as belonging to the same MLST, excluding single locus variants. After reference-based mapping, as described above, and assessment of intrahost pairwise SNP distance, a SNP cut-off of 100 was used to empirically delineate distinct intrahost strains, i.e., genomes diverging from the reference by more than the cut-off were considered a separate strain indicative of a distinct carriage event. Therefore, co-carriage was defined as individuals carrying more than one strain as defined by MLST type or SNP cutoff of 100 and mono-carriage as carrying one only strain. Throughout the text, the term strain is used to define genetically distinct SA isolates and intrahost isolates belonging to the same strain referred to as a population. The term sample is used to describe the swabs collected from a participant. Last, the term isolate or genome refer to an individual bacterium and have no connotation of genetic relatedness of population structure.

### Co-carriage dynamics

We evaluated individual and household risk factors for co-carriage using log-binomial regression to estimate prevalence ratios (PR) and 95% confidence intervals as previously described [24]. Given the small sample size, a limited multivariable analysis was performed, and only age-adjusted PRs are presented. SAS software, version 9.4 of the SAS System for Windows (SAS Institute), was used for the risk factor analysis.

Next, we assessed discordance in antibiotic resistance (*mecA*, *blaZ*, *mecA*, *AadD*, *Aph3.III*, *Sat*4A, *Spc*, *erm*, *mphC*, and *msrA*) and virulence factor (LukSF-PV, TSST, *etb*, Enterotoxin, *can*, *chp*, *scn*, and *vWbp*) determinants among co-carried strains, first considering co-carriage of MRSA and MSSA. We then identified variation in antibiotic resistance and virulence profiles, focusing on gain of determinants. For example, if an intrahost population was comprised of four isolates and one of them possessed *erm*, we considered that as discordance in antibiotic profile. We chose this approach because genotypic identification of determinants is more prone to false negatives than false positives due to genome assembly errors. In instances when differences of multiple determinants was identified, we further investigated differences in plasmid and bacteriophage content.

Last, we examined the phylogenetic relationship between co-carried strains. We first visualized the ML phylogeny in a Circos plot with connections between co-carried isolates belonging to the same participant. Then, we sought to determine whether co-carried strains were more likely to be closely related phylogenetically as compared to other non-co-carried strains. After abstracting a pairwise distance matrix from the ML phylogeny, we plotted the distribution of pairwise distances among co-carried strains and non-co-carried strains separately. To statistically test whether the distances among co-carried and non-co-carried strains were significantly different, we conducted a permutation test by subsampling 200 pairs of isolates equally balanced between co-carried and non-co-carried strains for 1000 iterations. We then conducted a Mann-Whitney U test comparing the two distributions of mean ML distances to determine if they were the same and to obtain an empirical p-value. To assess the effect size, we calculated Cohen’s D and defined small effect size as d=0.2, medium effect size as d=0.5, and large effect size as d=0.8 [39].

## RESULTS

### *Staphylococcus aureus* Carriage

Among the 60 adult participants, SA carriage positivity was similar by site: 36 (60.0%) in AN, 30 (50.0%) in NP, and 34 (56.7%) in OP (Figure 1 panel A). Twenty-eight (46.7%) participants carried at only one anatomical site, 24 (40.0%) participants carried at two anatomical sites, and 8 (13.3%) participants carried at three anatomical sites. The most common carriage site was OP (60.7%) among one-site carriers and AN+NP (62.5%) among two-site carriers. Between 4 and 9 isolates were sequenced for each participant (mean=5.2, standard deviation [SD]=1.6), for a total of 310 isolates across all participants (Table 1), including 108 (34.8%) from AN, 90 (29.0%) from NP, and 112 (36.1%) from OP.

**Figure 1.**
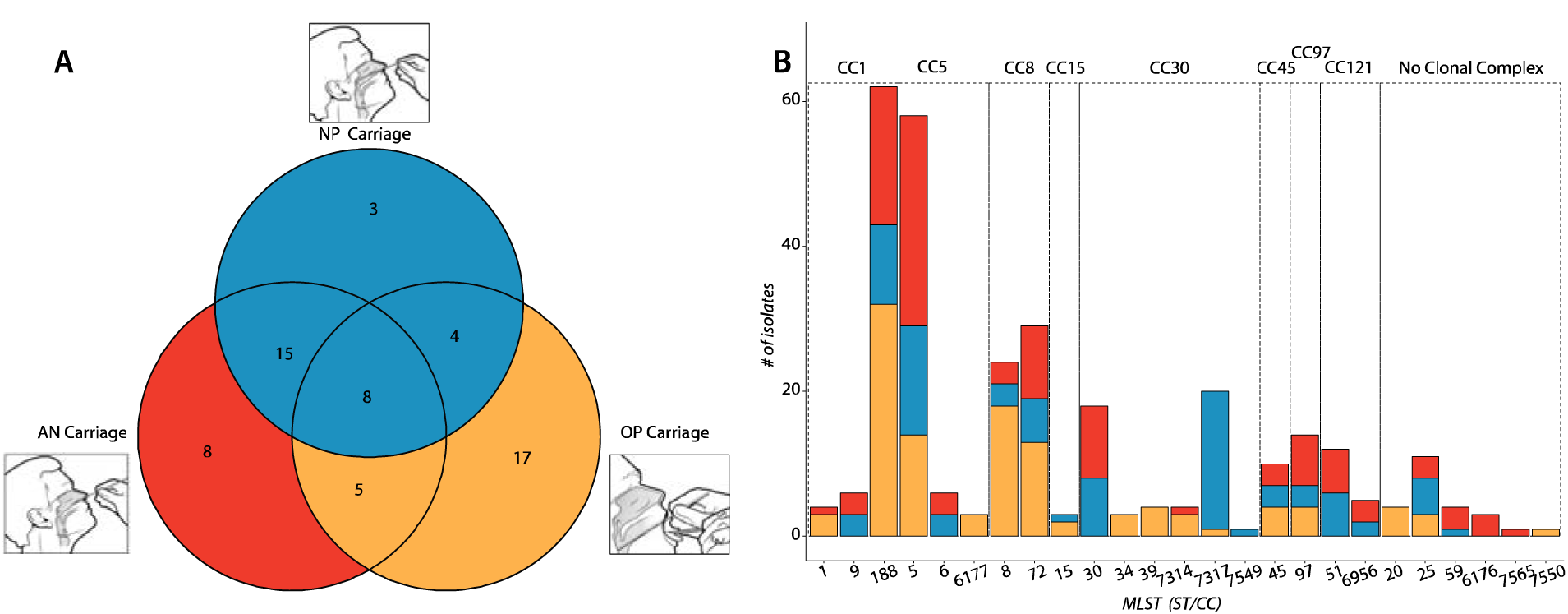
A. Staphylococcus aureus Carriage among Indigenous Adults. Each of the three circles represents an anatomical site of carriage and counts within denote the number of participants with carriage at the given sites***. B. Lineage of Isolates.*** Number of isolates belonging to each clonal complex (CC), and to each multi-locus sequence type (MLST) within each clonal complex is shown. Each bar is divided to indicate the number of isolates taken from each anatomical site; OP = oropharynx (yellow), NP = nasopharynx (blue), AN = anterior nares (red)

**Table 1:** *Staphylococcus aureus* Isolates by Anatomical Site among Indigenous Adults. Number of sequenced *S. aureus* isolates and proportion of total from each anatomical site, by whether they were taken from a participant that carried *S. aureus* at one, two, or three anatomical sites.

| Carriers | AN isolates<br>(% of total) | NP isolates<br>(% of total) | OP isolates<br>(% of total) | Total isolates<br>(% of total) |
| --- | --- | --- | --- | --- |
| 1-site carriers | 32 (10.3) | 15 (4.8) | 74 (23.9) | 121 (39.0) |
| 2-site carriers | 52 (16.8) | 51 (16.5) | 18 (5.8) | 121 (39.0) |
| 3-site carriers | 24 (7.7) | 24 (7.7) | 20 (6.5) | 68 (21.9) |
| **Grand Total** | **108 (34.8)** | **90 (29.0)** | **112 (36.1)** | **310** |

### Population Structure

To classify isolates by their major lineages, we determined the MLST of each SA genome. This identified 25 unique STs belonging to 8 major clonal complexes (CCs), led by CC1 (23.2%), CC5 (21.6%), CC8 (17.1%), and CC30 (16.1%) (Figure 1 panel B). The most prevalent CCs, CC1 and CC5, were both dominated by single MLSTs, ST188 and ST5, respectively. The distribution of CC by body site was significantly different. Notably, CC5 was disproportionately carried in the anterior nares, with 47.8% of CC5 isolates obtained from that anatomic site, CC30 was disproportionately carried in the nasopharynx, and CC1 and CC8 were more frequently isolated from the oropharynx (Table 2). Similarly, within CCs, some MLSTs were dominated by isolates from specific anatomical sites. For example, 95.0% of ST7317 isolates were obtained from the nasopharynx. Assessing co-carriage, we found that 35 (58.3%) participants carried isolates belonging to a single MLST, 19 (31.7%) carried two MLST, and 6 (10.0%) carried three MLST, delineating 91 distinct populations among 60 individuals based on MLST and giving an overall proportion of co-carriage with multiple MLSTs of 41.7% (25/60).

**Table 2.** Clonal complex (CC) prevalence comparison among body sites: anterior nares (AN), oropharynx (OP), and nasopharynx (NP). Pairwise comparisons of carriage by anatomic site. Odd ratios and 95% highest posterior density (HPD) are given for each comparison, and significant values are bolded.

| Clonal Complex | Anterior nares (AN, n=108) vs Other (n=202) |  | Nasopharynx (NP, n=90) vs Other (n=220) |  | Oropharynx (OP, n=112) vs Other (n=198) |  |
| --- | --- | --- | --- | --- | --- | --- |
|  | Body Site | Odds Ratio (95% HPD) | Body Site | Odds Ratio (95% HPD) | Body Site | Odds Ratio (95% HPD) |
| CC1 | AN (n=23) | 0.87 (0.63-1.18) | NP (n=14) | 0.59 (0.42-0.80) | OP (n=35) | **1.69 (1.29-2.41)** |
|  | Other (n=49) | Ref | Other (n=58) | Ref | Other (n=37) | Ref |
| CC5 | AN (n=32) | **1.68 (1.21-2.34)** | NP (n=18) | 0.91 (0.66-1.24) | OP (n=17) | 0.60 (0.42-0.83) |
|  | Other (n=35) | Ref | Other (n=49) | Ref | Other (n=50) | Ref |
| CC8 | AN (n=13) | 0.59 (0.37-0.88) | NP (n=9) | 0.51 (0.32-0.79) | OP (n=31) | **2.54 (1.76-3.72)** |
|  | Other (n=40) | Ref | Other (n=44) | Ref | Other (n=22) | Ref |
| CC30 | AN (n=11) | 0.52 (0.32-0.78) | NP (n=28) | **3.15 (2.18-4.69)** | OP (n=11) | 0.51 (0.33-0.76) |
|  | Other (n=39) | Ref | Other (n=22) | Ref | Other (n=39) | Ref |
| CC45 | AN (n=3) | 0.78 (0.22-2.00) | NP (n=3) | 1.11 (0.38-2.60) | OP (n=4) | 1.22 (0.46-3.33) |
|  | Other (n=7) | Ref | Other (n=7) | Ref | Other (n=6) | Ref |
| CC97 | AN (n=7) | 1.86 (0.88-4.67) | NP (n=3) | 0.67 (0.25-1.44) | OP (n=4) | 0.70 (0.29-1.71) |
|  | Other (n=7) | Ref | Other (n=11) | Ref | Other (n=10) | Ref |
| CC121 | AN (n=9) | **2.00 (1.09-4.75)** | NP (n=8) | **2.11 (1.13-5.25)** | - |  |
|  | Other (n=8) | Ref | Other (n=9) | Ref |  |  |

### Intrahost diversity and co-carriage

To obtain greater resolution of intrahost diversity and co-carriage, we assessed pairwise SNP distances (Figure 2). Of the 91 populations delineated by MLST, 78 had more than one isolate, allowing for pairwise comparison. Plotting intrahost pairwise distances for these populations showed that most resolved into genomically cohesive groups of isolates with distances ranging from 0-100 SNPs and some isolates forming a long tail distribution from 450-750 SNPs (Figure 2). As mean pairwise distances are often used to describe population diversity, we additionally plotted the mean intrahost distances for each population, finding that most distances fell into the 0-50 SNPs range with a tail at 300-500 SNPs (mean = 25.02, SD=95.05) (Supplemental Figure 1). Using a cut-off of 100 SNPs, we identified four participants that had intrahost populations belonging to the same MLST but comprised of divergent isolates indicative of distinct populations. These isolates differed by hundreds of SNPs (mean= 618.2) and did not have evidence of intrahost recombination events based on the examination of the spatial distribution of SNPs in the genome. All four participants had also been identified as co-carriers based on MLST.

**Figure 2.**
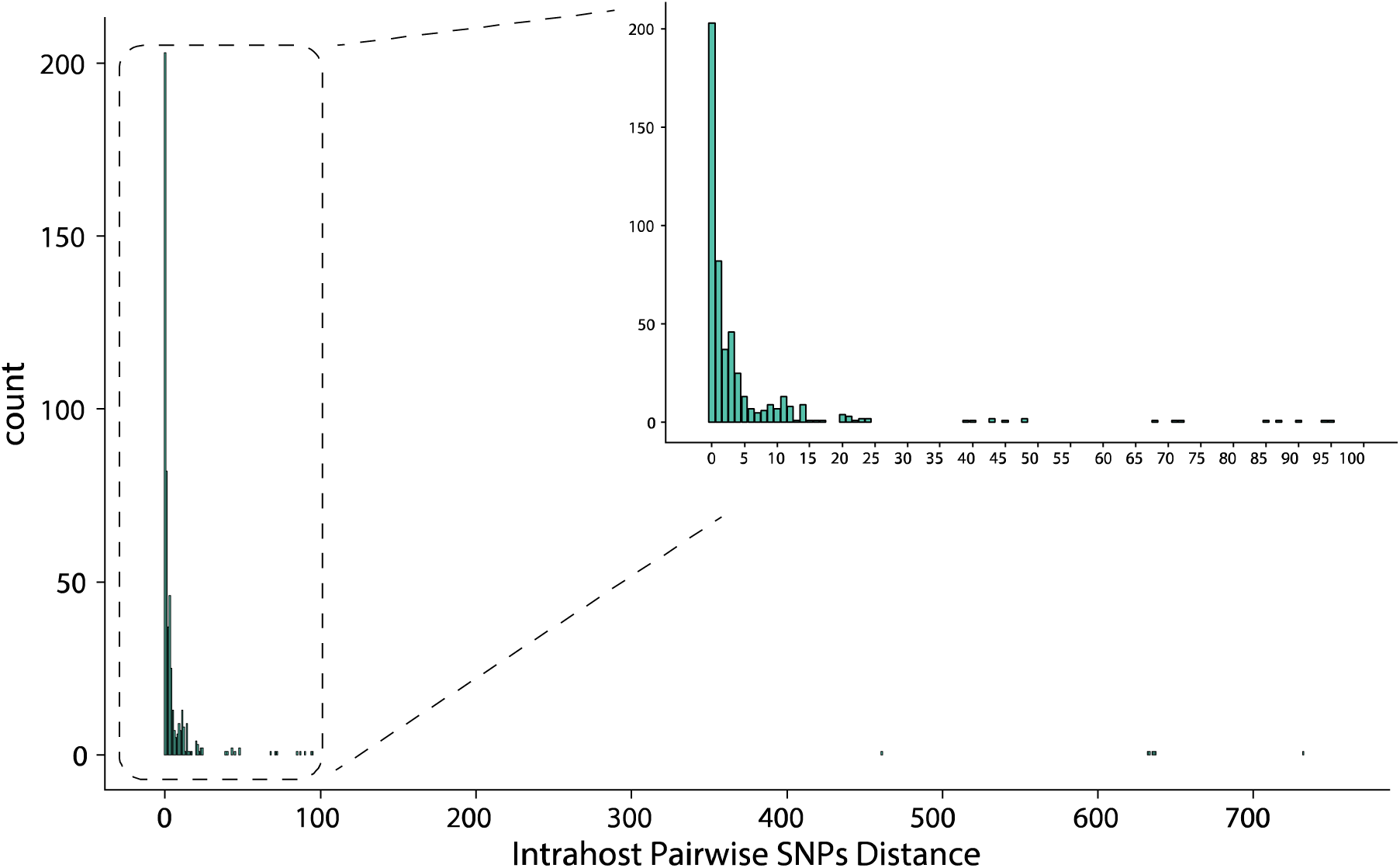
Pairwise SNP-distances among intrahost individual SA populations with ≥2 isolates of the same MLST (n=78). An enlarged view of pairwise distances in the 0-100 SNP region of interest is shown on the top right of the figure. Several distances fall above the 39 SNP cutoff often used for inferring an epidemiological linkage.

Taken together, using WGS data, we identified 95 distinct SA populations among 60 participants and an overall proportion of co-carriage of 25/60 (41.7%). If the 6 participants excluded for low numbers of isolates were single population carriers, the overall proportion of co-carriage would be 33.3% (25/66). Further, while WGS data did not identify additional co-carriers, it revealed that two participants each carried four distinct populations and two carried three. Of note, using a more conservative cut-off of 39 SNPs, as previously suggested by Hall et al. [10], we would have delineated four additional co-carriers. Among co-carriage instances, 13/25 (52.0%) involved strain variation within a specific anatomic site and 12 instances involved distinct strains compartmentalized at an anatomic site. In addition, 14, 18, and 20 instances of co-carriage involved OP, NP, and AN sites, respectively. Notably, of the 18 instances of co-carriage involving the NP, 7 involved the same strain found at both the AN and NP site.

**Supplemental Figure 1.**
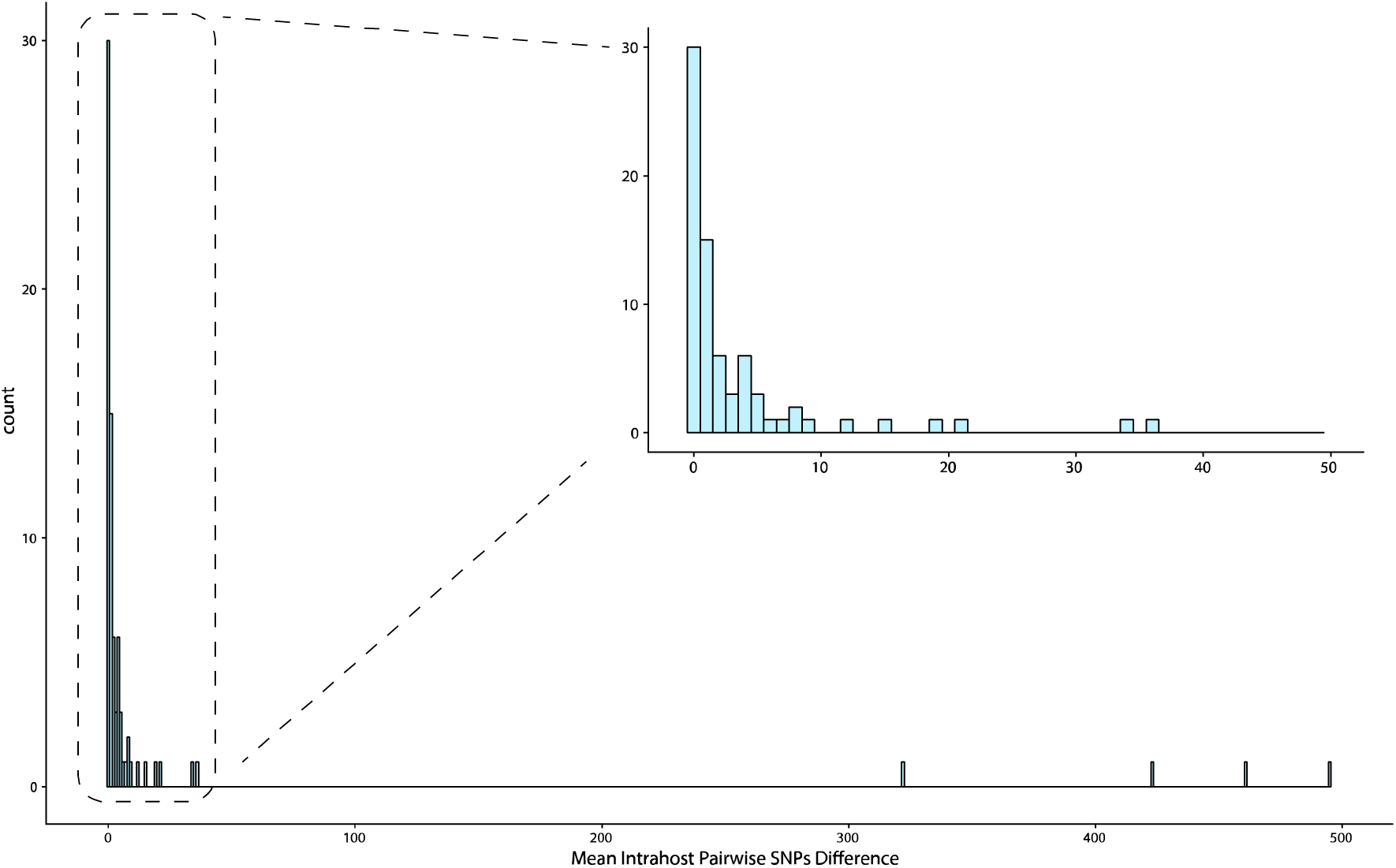
Mean pairwise SNP-distances among intrahost individual SA populations with ≥2 isolates of the same MLST (n=78). An enlarged view of pairwise distances in the 0-50 SNP region of interest is shown on the top right of the figure.

### Risk factors for co-carriage

Individual (Table 3) and household (Table 4) risk factors for co-carriage of distinct SA populations were assessed. Hospitalization in the last 6 months, gym use in the last 6 months, and living in a household with seven or more people were significantly correlated with co-carriage in bivariate and age-adjusted analyses. While participation in team sports and activities in the last 6 months and having an average of more than two people per bedroom were associated with co-carriage in bivariate analysis, these results were no longer statistically significant after adjusting for age. The three participants with co-carriage who reported large household size, also had recent healthcare exposure, indicating that multiple risk factors may contribute to high rates of co-carriage.

**Table 3.** Individual characteristics of study participants and correlates of co-carriage among Indigenous adults in the Southwest US in 2017.

|  |  | Total population<br>(N=60)<br>n (%) | Co-carrier<br>(n=25)<br>n (%) | p-value<sup>a</sup> | Crude prevalence ratio<br>(95% CI) | Age-adjusted prevalence ratio<br>(95% CI) |
| --- | --- | --- | --- | --- | --- | --- |
| Age group<br>(years) | 18-39 | 23 (38.3) | 13 (56.5) | 0.21 | Ref | - |
|  | 40-64 | 23 (38.3) | 7 (30.4) |  | 0.54 (0.26, 1.10) | - |
| | $\geq 65$ | 14 (23.3) | 5 (35.7) | | 0.63 (0.29, 1.39) | - |
| Sex | Male | 12 (20.0) | 7 (58.3) | 0.21 | 1.56 (0.85, 2.84) | 1.37 (0.79, 2.39) |
|  | Female | 48 (80.0) | 18 (37.5) |  | Ref | Ref |
| Type of living facility | Residence | 60 (100.0) | 25 (41.7) | - | Ref | Ref |
|  | Currently homeless | 0 | 0 |  | - | - |
| Times likely to bathe or shower in a typical week | 1-2 | 9 (15.3) | 3 (33.3) | 0.39 | Ref | Ref |
|  | 3-4 | 18 (30.5) | 10 (55.6) |  | 1.67 (0.61, 4.59) | 1.47 (0.50, 4.35) |
|  | 5-6 | 6 (10.2) | 1 (16.7) |  | 0.50 (0.07, 3.75) | 0.43 (0.05, 3.79) |
|  | 7+ | 26 (44.1) | 11 (42.3) |  | 1.27 (0.45, 3.55) | 1.16 (0.39, 3.42) |
| Current tobacco smoker | No | 51 (85.0) | 21 (41.2) | 1.00 | Ref | Ref |
|  | Yes | 9 (15.0) | 4 (44.4) |  | 1.08 (0.48, 2.40) | 0.91 (0.44, 1.87) |
| Current chewing tobacco user | No | 55 (91.7) | 24 (43.6) | 0.39 | Ref | Ref |
|  | Yes | 5 (8.3) | 1 (20.0) |  | 0.46 (0.08, 2.71) | 0.44 (0.08, 2.33) |
| Self-reported history of skin infections or complications (e.g. boils, spider bites, eczema) | No | 49 (81.7) | 20 (40.8) | 1.00 | Ref | Ref |
|  | Yes | 11 (18.3) | 5 (45.5) |  | 1.11 (0.54, 2.31) | 0.97 (0.48, 1.99) |
| Documented SA infection in the past 5 years | No | 58 (96.7) | 24 (41.4) | 1.00 | Ref | Ref |
|  | Yes | 2 (3.3) | 1 (50.0) |  | 1.21 (0.29, 5.00) | 1.75 (0.37, 8.18) |
| Antibiotic use in the past 14 days | No | 56 (96.6) | 24 (42.9) | 0.51 | Ref | Ref |
|  | Yes | 2 (3.5) | 0 |  | - | - |
| Any underlying health condition | No | 21 (35.0) | 12 (57.1) | 0.10 | Ref | Ref |
|  | Yes | 39 (65.0) | 13 (33.3) |  | 0.58 (0.33, 1.04) | 0.65 (0.36, 1.16) |
| Diabetes | No | 42 (70.0) | 19 (45.2) | 0.57 | Ref | Ref |
|  | Yes | 18 (30.0) | 6 (33.3) |  | 0.74 (0.35, 1.53) | 0.98 (0.43, 2.25) |
| Obesity | No | 28 (46.7) | 13 (46.4) | 0.48 | Ref | Ref |
|  | Yes | 32 (53.3) | 12 (37.5) |  | 0.81 (0.44, 1.47) | 0.86 (0.48, 1.54) |
| Behaviors in the past 6 months |  |  |  |  |  |  |
| Hospitalization | No | 51 (85.0) | 18 (35.3) | 0.03 | Ref | Ref |
|  | Yes | 9 (15.0) | 7 (77.8) |  | **2.20 (1.32, 3.67)** | **2.13 (1.20, 3.77)** |
| Surgery | No | 52 (86.7) | 20 (38.5) | 0.26 | Ref | Ref |
|  | Yes | 8 (13.3) | 5 (62.5) |  | 1.63 (0.86, 3.07) | 1.58 (0.91, 2.76) |
| Shared a towel | No | 46 (76.7) | 20 (43.5) | 0.76 | Ref | Ref |
|  | Yes | 14 (23.3) | 5 (35.7) |  | 0.82 (0.38, 1.79) | 0.97 (0.42, 2.27) |
| Shared a bed | No | 35 (58.3) | 13 (37.1) | 0.40 | Ref | Ref |
|  | Yes | 25 (41.7) | 12 (48.0) |  | 1.29 (0.71, 2.34) | 1.29 (0.71, 2.34) |
| Worn clothes more than once without washing | No | 39 (65.0) | 17 (43.6) | 0.79 | Ref | Ref |
|  | Yes | 21 (35.0) | 21 (35.0) |  | 0.87 (0.46, 1.68) | 0.88 (0.47, 1.65) |
| Shared clothes | No | 59 (98.3) | 25 (42.4) | 1.00 | Ref | Ref |
|  | Yes | 1 (1.7) | 0 |  | - | - |
| Gym use | No | 51 (85.0) | 19 (37.3) | 0.15 | Ref | Ref |
|  | Yes | 9 (15.0) | 6 (66.7) |  | **1.79 (1.00, 3.21)** | **1.84 (1.07, 3.17)** |
| Used a locker room | No | 56 (93.3) | 23 (41.1) | 1.00 | Ref | Ref |
|  | Yes | 4 (6.7) | 2 (50.0) |  | 1.22 (0.44, 3.41) | 1.00 (0.39, 2.54) |
| Went to a sweat lodge | No | 59 (98.3) | 24 (40.7) | 0.42 | Ref | Ref |
|  | Yes | 1 (1.7) | 1 (100.0) |  | - | - |
| Participated in organized team sports or activities | No | 56 (93.3) | 22 (39.3) | 0.30 | Ref | Ref |
|  | Yes | 4 (6.7) | 3 (75.0) |  | **1.96 (1.02, 3.79)** | 1.64 (0.98, 2.75) |
| Received any artistic (body-art) or cosmetic tattoos | No | 55 (91.7) | 23 (41.8) | 1.00 | Ref | Ref |
|  | Yes | 5 (8.3) | 2 (40.0) |  | 0.98 (0.32, 3.01) | 0.88 (0.33, 2.31) |
| Received any body or facial piercings | No | 57 (95.0) | 24 (42.1) | 1.00 | Ref | Ref |
|  | Yes | 3 (5.0) | 1 (33.3) |  | 0.81 (0.16, 4.15) | 0.70 (0.16, 3.09) |
| Used drugs for non-medical purposes | No | 57 (95.0) | 25 (43.9) | 0.26 | Ref | Ref |
|  | Yes | 3 (5.0) | 0 |  | - | - |

<sup>a</sup> p-value from Chi-square test or Fisher's exact test, as appropriate
Bold indicates $p < 0.05$ from Poisson regression with robust variance estimation

**Table 4.**
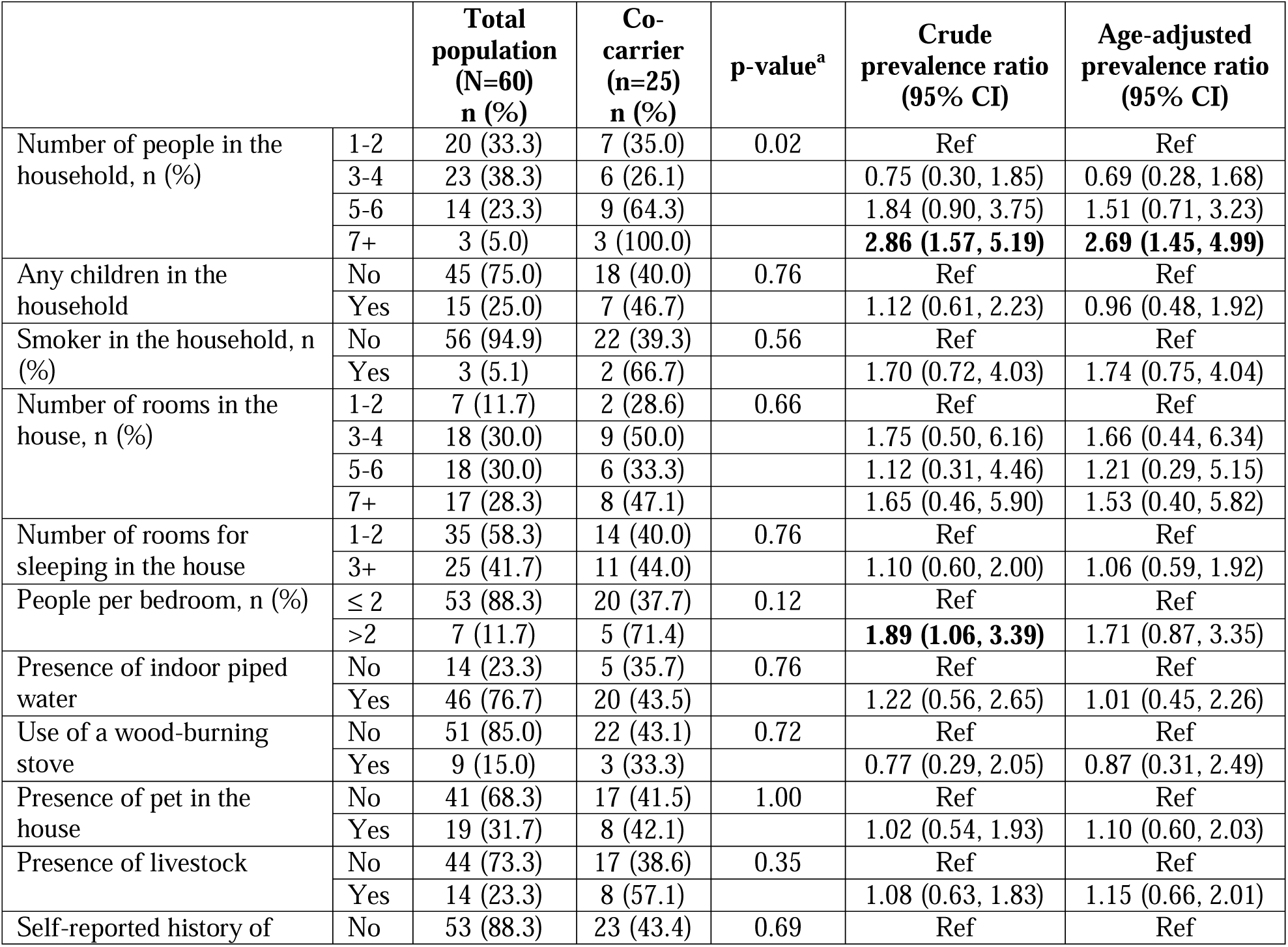
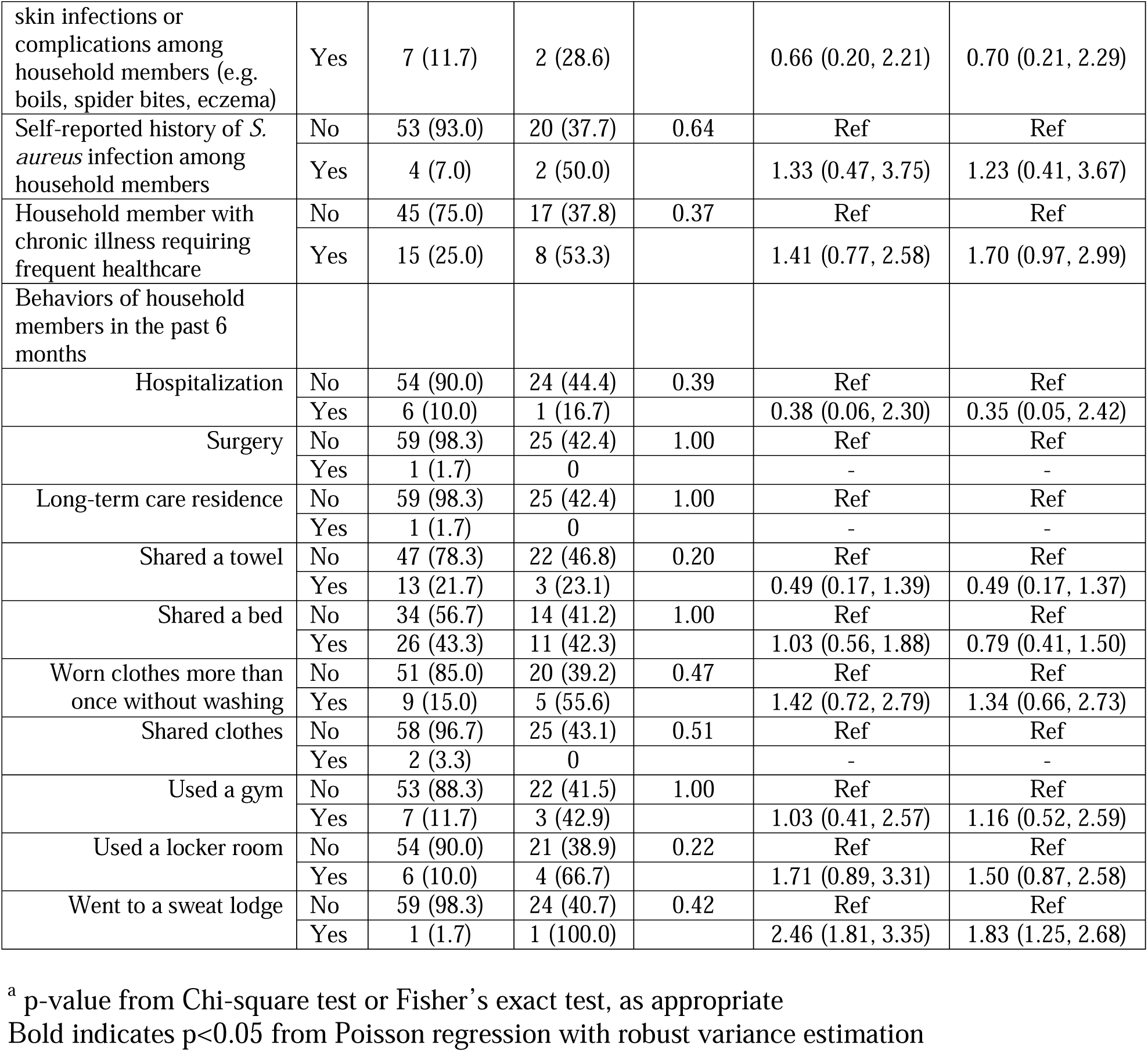
Household characteristics of study participants and correlates of co-carriage among Indigenous adults in the Southwest US in 2017.

### Phylogenomics

To visualize the population structure and phylogenetic relationship between isolates, we constructed a maximum-likelihood phylogeny of all 310 genome assemblies and mapped the distribution of antibiotic resistance and virulence determinants (Figure 3). The phylogeny shows that the majority of isolates cluster into 8 major clades corresponding to CC. MRSA isolates, denoted by a star tip point, are almost exclusively found in CC8, which also predominantly harbored strains demonstrating aminoglycoside and macrolide resistance determinants and LukSF-PV encoding Panton-Valentine Leucocidin. We found 25 instances of intrahost variation in antibiotic resistance determinants, accounting for 41.7% of the 60 participants studied. Of these, 20 instances were among individuals identified as co-carriers while the other 5 instances were among mono-carriers. In three instances, we identified co-carriage of MRSA and MSSA strains belonging to two different MLSTs. The most common variation was observed with *blaZ* (18/25, 72.0%), which encodes β-lactamase and is associated with penicillin non-susceptibility, followed by *fosB* (10/25, 40.0%) and *erm* (8/25, 32.0%), which confer fosfomycin and macrolide resistance, respectively. Regarding variation in virulence determinants, we identified 38 instances of intrahost variation among 34 participants. Of these, 25 instances were among individuals identified as co-carriers, nine instances were among mono-carriers, and four instances were among individuals with variation among co-carried strains (i.e., same MLSTs but a distinct population based on SNP distances) as well as within a strain/population. Finally, to investigate the phylogenetic relationship among co-carried strains, we visualized the population structure illustrating links between strains isolated from the same participant and plotting the distribution of ML pairwise distances (Figure 4). While the pairwise distances among co-carried strains were significantly less (i.e., more closely related) than the overall pairwise distances among non-co-carried strains, the effect size was small (Supplemental Figure 2).

**Figure 3.**
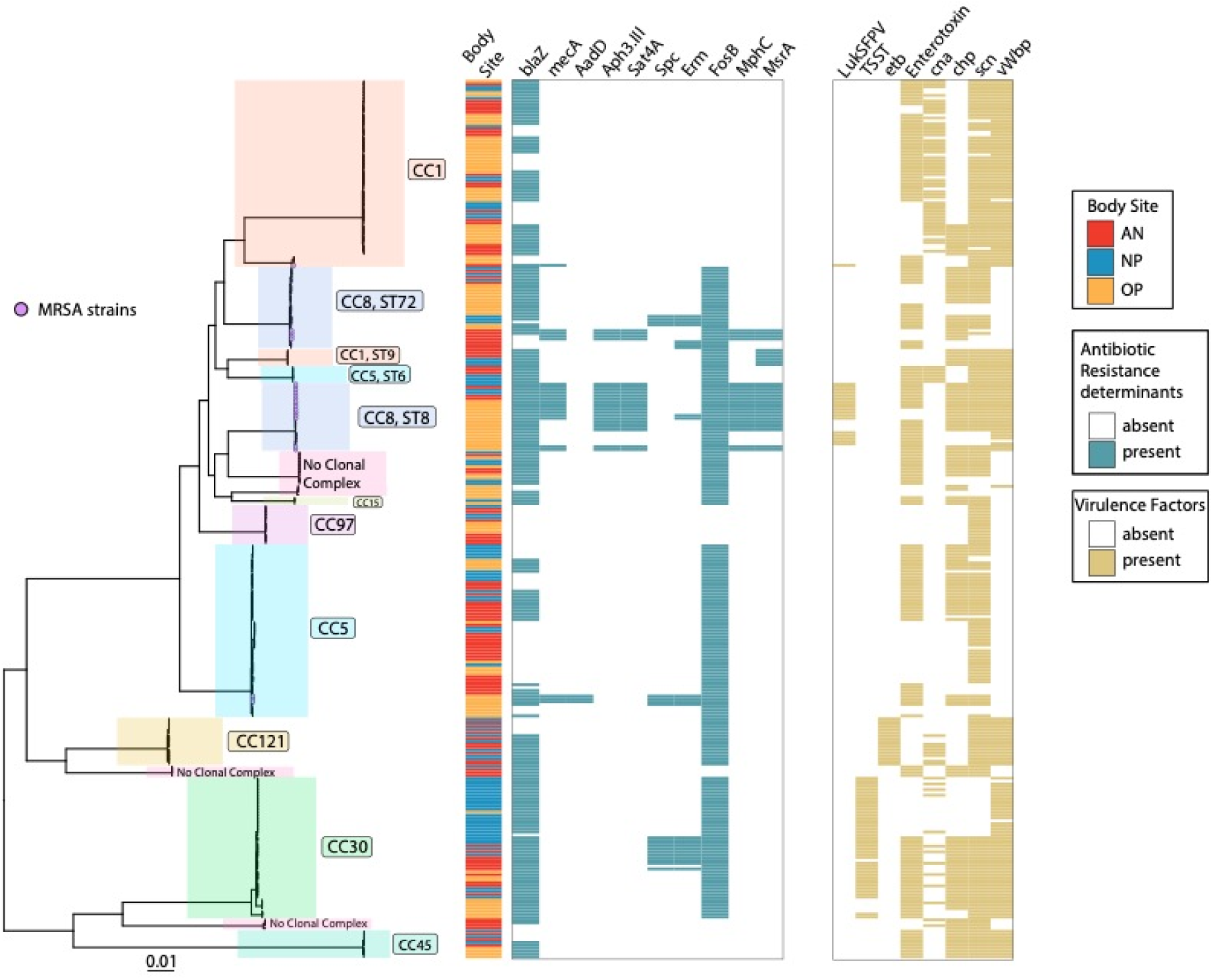
Maximum-likelihood phylogeny of carried SA isolates and their associated traits. A maximum-likelihood phylogeny was generated from a core-SNP alignment of all 310 isolates. Each tip corresponds to an isolate; a purple circle at the tip indicates the presence of *mecA* conferring methicillin resistance. Clades are organized by clonal complex. To the right of the tree, presence/absence matrices of antibiotic resistance determinants and virulence factors are shown. Antibiotic resistance determinants in order: *blaZ*, *mecA*, *AadD*, *Aph3.III*, *Sat*4A, *Spc*, *erm*, *mphC*, and *msrA*. Virulence factors in order: LukSF-PV, TSST, *etb*, Enterotoxin, *can*, *chp*, *scn*, and *vWbp*.

**Figure 4:**
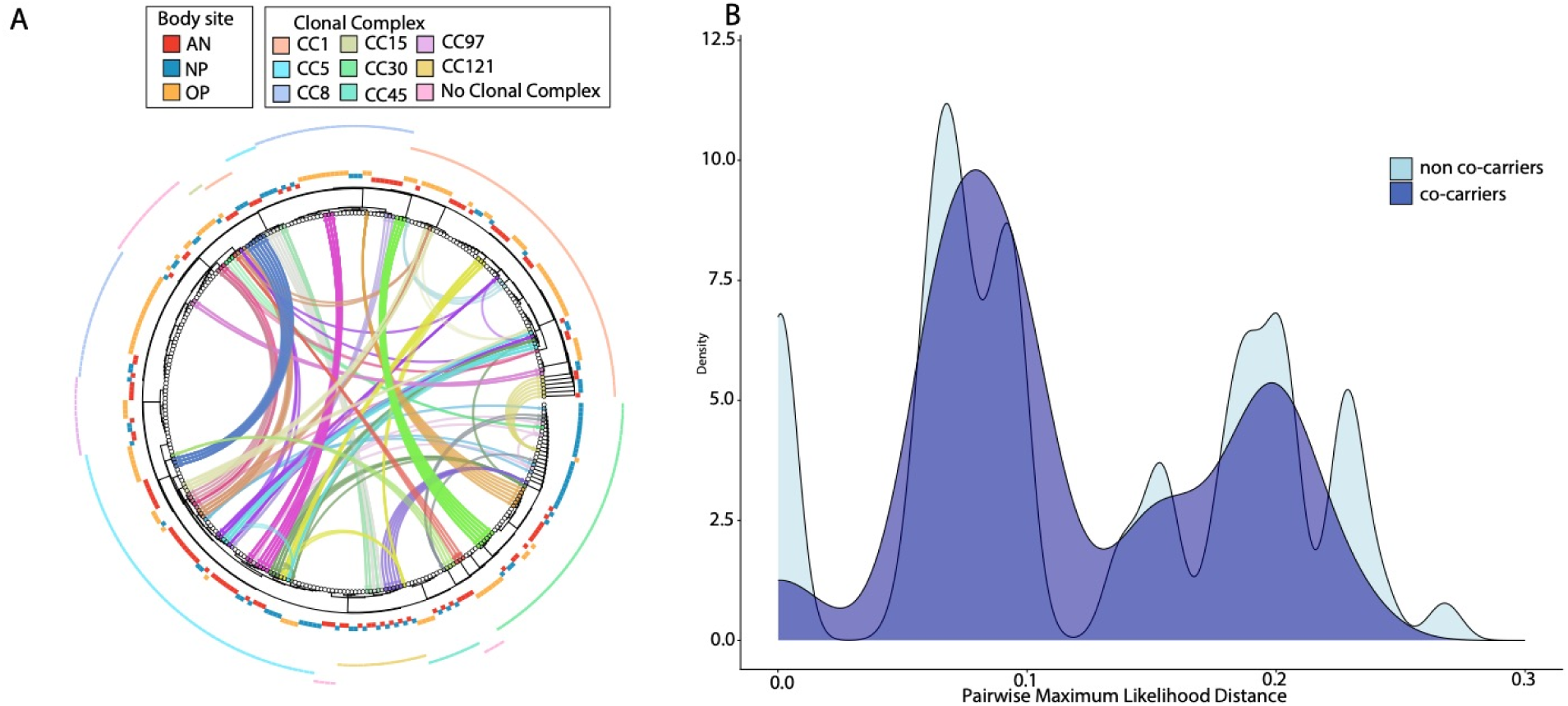
Circos plot of maximum-likelihood (ML) phylogeny and the relationship between co-carried SA strains. A) A ML phylogeny was generated from a core-SNP alignment of all 310 isolates. Each tip corresponds to an isolate. The first ring around the tree denotes anatomical site: AN=anterior nares, NP=nasopharynx, or OP=oropharynx. The outermost ring around the tree denotes clonal complex. Inside the tree, connections are drawn between isolates that belong to the same participant; each participant is represented by a unique color. B) Density histogram plot comparing the pairwise ML genetic distance between co-carried strains from the same individual (dark blue) and the overall distribution of pairwise distances among all strains from different individuals (non co-carried strains, light blue).

**Supplemental Figure 2:**
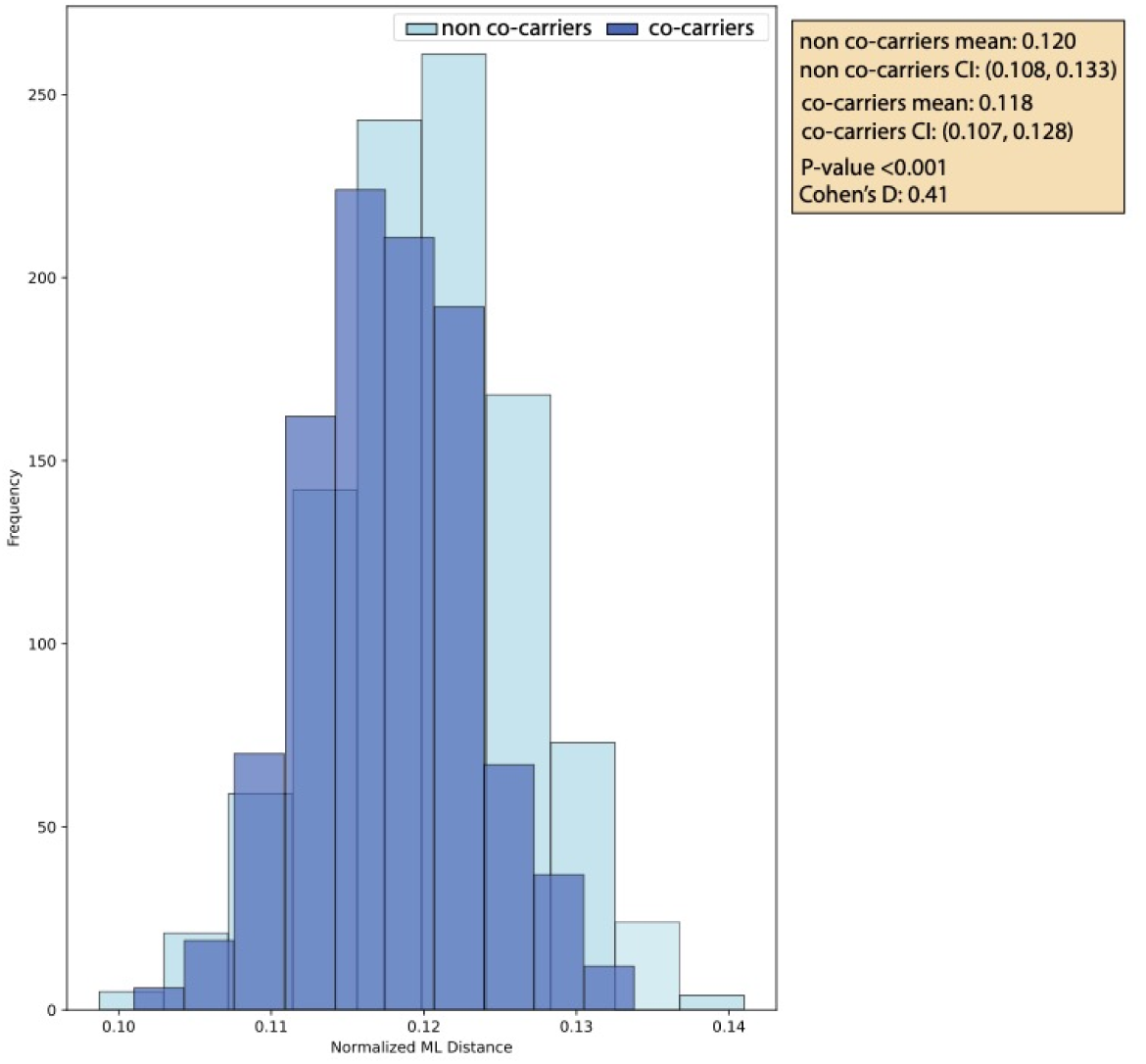
Results of permutation test assessing the maximum likelihood (ML) distances among co-carried from the same individual and non-co-carried strains from all individuals. Pairwise ML distances were subsampled to obtain 200 pairs of isolates equally balanced between co-carried and non-co-carried strains for 1000 iterations. Empirical p-value results are for the Mann-Whitney U test comparing the two distributions of mean distances. The Cohen’s D value represents the effect size defined as small d=0.2, medium d=0.5, and large d=0.8.

## DISCUSSION

Our investigation of co-carriage and intrahost diversity of SA among Indigenous adults in the Southwest US, which involved sampling multiple respiratory anatomical sites and genomic analysis of multiple isolates, revealed a co-carriage prevalence of 41.7%. Notably, we resolved the degree of co-carriage (i.e., the number of distinct strains carried), finding six individuals carrying three distinct strains and two carrying four strains. Further, intrahost populations were found to vary in genome content, including antibiotic resistance and virulence determinants, which has considerable implications for carriage studies. Overall, our investigation of the intrahost diversity of SA carriage using a multi-site, multi-isolate, bacterial genome sequencing approach addressed previous limitations and updates our understanding of intrahost diversity during carriage.

Historically, SA carriage was generally thought to be clonal with only one strain present [40]. Few studies have systematically explored co-carriage of multiple SA strains. An early study of food handlers in 2003 revealed 78.6% of participants (n=14) carried more than one strain, as defined by PFGE [17]. Subsequently, a comprehensive study of the clonality of SA carriage found that 14 out of 148 (9.4%) subjects carried isolates of different PFGE profile, and 7 of these 14 subjects carried more than one strain as defined by MLST [18]. A pediatric study reported that 30.4% of carriers were colonized with more than one genotype as defined by multiple-locus variable-number tandem repeat fingerprinting [19], and Votintseva *et al*. found that over a 24- month period, 18% of subjects were co-colonized at some point by more than one strain as defined by *spa*-type [20].

More recently, the advent of bacterial genome sequencing demonstrated that previous typing methods lacked the resolution to delineate distinct strains and within host populations. Here, MLST was able to identify most co-carriers; however, the degree of co-carriage was only fully resolved through dense genomic sampling of multiple isolates from three anatomical sites. Our findings have significant implications for the use of genomic data to track pathogen transmission as well as understanding intra- and inter-host evolutionary rates. For example, previous studies have sought to establish a straightforward SNP-based cut-off to identify epidemiological linkage with upper-end limits set to 40 SNPs based on an estimated rate of 8 mutations per genome per year [10,41]. However, bioinformatics methods can vary considerably among studies, resulting in measurement differences in SNP distances. In particular, genome assembly methods and reference selection [42], in the case of reference-based assembly, can impact the “callable” portions of the genome, thereby reducing resolution and artificially reducing distances. Use of an internal reference generated through hybrid assembly of an intrahost strain increased our ability to assess microevolution at an unmatched level. Yet it complicates our determination of intrahost populations as we have limited knowledge of the expected genomic variation when multiple upper respiratory sites are densely sampled. As a result, delineating populations that include strains with distances ranging from 40-100 SNPs remains speculative as these outliers could result from large transmission bottleneck size, extended durations of carriage, incomplete purifying selection, repeated self-inoculation from a reservoir body site, or recurrent/ongoing household transmission [43]. The latter is consistent with the observed association between household size and co-carriage, further supporting the concept of long-term ongoing transmission of an endemic strain within the household. Subsequent longitudinal studies with repeated sampling of the household would allow us to resolve these dynamics. Nevertheless, our findings suggest that a high density of sampling at multiple body sites is important for transmission studies.

The oropharynx and anterior nares are the most frequently sampled sites for SA carriage [44,45]. While the nasopharynx is rarely sampled for SA [46] and is not thought to be a primary carriage site, we found that 50% of adult SA carriers had nasopharyngeal colonization. While it is possible that passage of a swab through the anterior nares during nasopharyngeal sampling could result in “contamination” of the sample, the repeated identification of nasopharyngeal SA populations distinct from oropharyngeal and/or anterior nares samples co-carried in the same individual suggests these findings represent true nasopharyngeal carriage. In addition, we found evidence of an association between specific clonal complexes and anatomic site of isolation.

CC5 was statistically significantly associated with isolation from the anterior nares, CC30 from the nasopharynx, and CC1 and CC8 from the oropharynx. ST7317, a novel MLST belonging to CC30, was comprised almost entirely of isolates from the nasopharynx. While previous studies have observed an association of specific lineages (PFGE-types of STs) with disease conditions such as ocular infections, atopic dermatitis, and toxic shock syndrome [47–49], there are limited data on variation in strain type by anatomical site of carriage. A previous finding that PFGE-type USA300 (ST8) was associated with a high prevalence of oropharyngeal carriage in a New York prison population remains one of the few examples of preferential carriage by anatomic site and is consistent with our findings here [50]. Overall, our findings strongly suggests that lineages may be adapted for colonization of specific anatomical sites, or that repeated exposure to new strains may compartmentalize populations across sites. These ideas require further exploration in future studies.

SA strains can vary considerably in genome content due to horizontal gene transfer [51], and transfer of plasmids and bacteriophages has been documented during co-colonization [52]. We observed variation in antibiotic and virulence determinants throughout the phylogeny of co-carried strains. Assessing intrahost differences in more detail, we found that an appreciable proportion of individuals with both co- and mono-carriage had clinically relevant differences in virulence and antibiotic resistance determinants, even in instances where the individual populations resided in different anatomical sites. Most notable were instances of co-carriage of MRSA and MSSA strains, macrolide resistance conferred my *erm*, and virulence genes for exfoliative toxin (*etb/eta*) and toxic-shock syndrome. Indeed, co-carriage provides an ideal setting for the exchange of mobile genetic elements. This finding further supports the need to sample multiple body sites in epidemiologic studies or clinical MRSA surveillance, as sampling a single anatomical site may miss MRSA carriage.

Further, little is known about intraspecies interactions during co-carriage. Co-carried strains are in a competition for resources while simultaneously evading the host immune system and other colonizing species. Previously, competitive exclusion between strains was thought to limit the incidence of co-carriage [53]. For example, carriage of MSSA at a single anatomical site was found to be protective for MRSA acquisition [54]. As such, we posited that co-carried strains would be genomically divergent, since this would potentially maximize the antigenic divergence, thereby avoiding any strain specific immunity to superantigens or capsular polysaccharide [55]. However, we observed no overt association between co-carried strains based on genetic divergence, as illustrated by the complexity in the Circos plot and distribution of pairwise SNP distances. Notwithstanding, an increased tolerance of a strain for co-colonization could be an evolutionary advantage as it would provide an opportunity for onward transmission as well as acquisition of mobile elements. Further, niche expansion through the ability to inhabit multiple respiratory sites would avoid direct competition for binding to epithelial cells and would further explain why we frequently observed compartmentalization of distinct population across anatomical sites. A full accounting of genome content variation between mono- and co-carried strains as well as phenotypic and competition experiments may further elucidate the existence of preferred strain pairs and how these dynamics relate to strain prevalence and carriage duration.

Crowding, participation in team sports, and healthcare contact are previously recognized risk factors for community-associated SA carriage and disease [56]. Therefore, one might expect these same risk factors to increase the likelihood of co-carriage. Indeed, we observed healthcare, household, and community risk factors associated with carriage of multiple SA strains. In our previous analysis, several household risk factors were associated with carriage, including history of SA infection among household members and household members sharing a bed or using a gym or locker room [23]. Consistent with these previous findings, number of people in the household and number of people per bedroom, both proxies for crowding, and healthcare exposure were found to be significantly associated with increased likelihood of co-carriage. As household size increases, multiple individual risk factors are compounded, likely leading to ongoing transmission of “endemic” household strains and increasing the potential for introduction of new strains acquired from outside the household (e.g., gym or hospital). Together, this underscores the significance of household transmission of SA, as has been reported [57,58], and suggests an opportunity for interventions such as decolonization, enhanced environmental cleaning, or education campaigns.

Our study was not without limitations; primarily, the inclusion only of adults leaves intrahost SA carriage in children yet to be explored. As children are an important reservoir for respiratory pathogens and contribute to community transmission [59], future study is merited. The small sample size limited our ability to fully explore the individual and household risk factors associated with co-carriage. Additionally, this study was cross-sectional, assessing samples taken only at a single timepoint for each participant. Future work should include longitudinal sampling, to explore the intrahost dynamics of SA carriage over time and impact of co-carriage on the duration of carriage.

Overall, this work offers a high-resolution analysis of intrahost diversity among SA carriers. We found that an appreciable proportion of carriers were colonized by more than one MLST. In particular, our novel use of hybrid genome sequencing to generate an intrahost reference allowed us to assess intrahost evolution at a previously unexplored level of resolution. Together, these findings indicate that the diversity of SA within carriers is greater than previously appreciated. As accurate identification and characterization of SA carriage is central to surveillance activities and genomic epidemiology studies, we show the importance of considering intrahost diversity including co-carriage and incorporating these methods into future work.

## Data Availability

All data produced in the present study are available upon reasonable request to the authors.

