## Supplementary figures and images for "Multi-strain carriage and intrahost diversity of *Staphylococcus aureus* among Indigenous adults in the US"

### Supplemental Figure 1

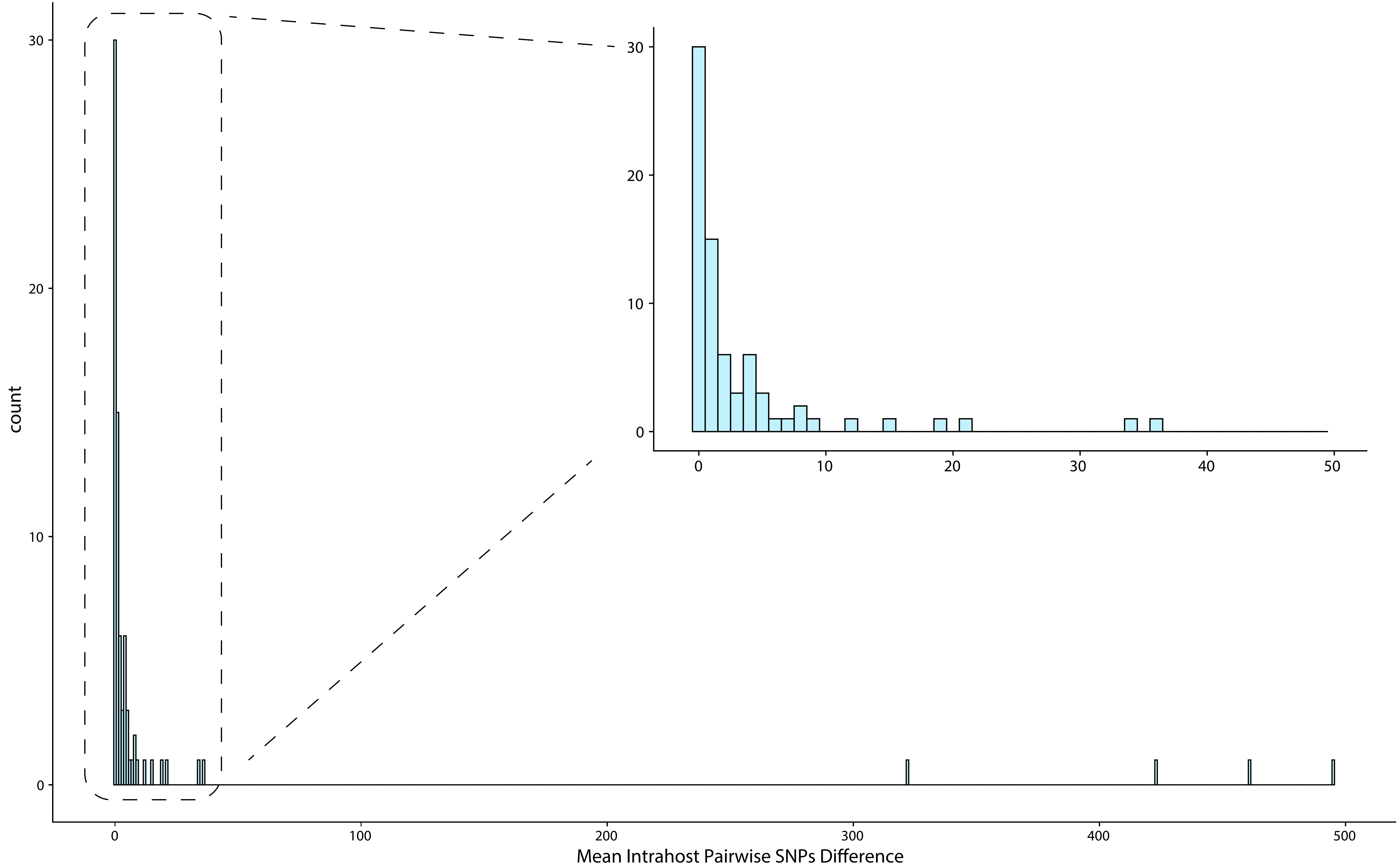

### Supplemental Figure 2

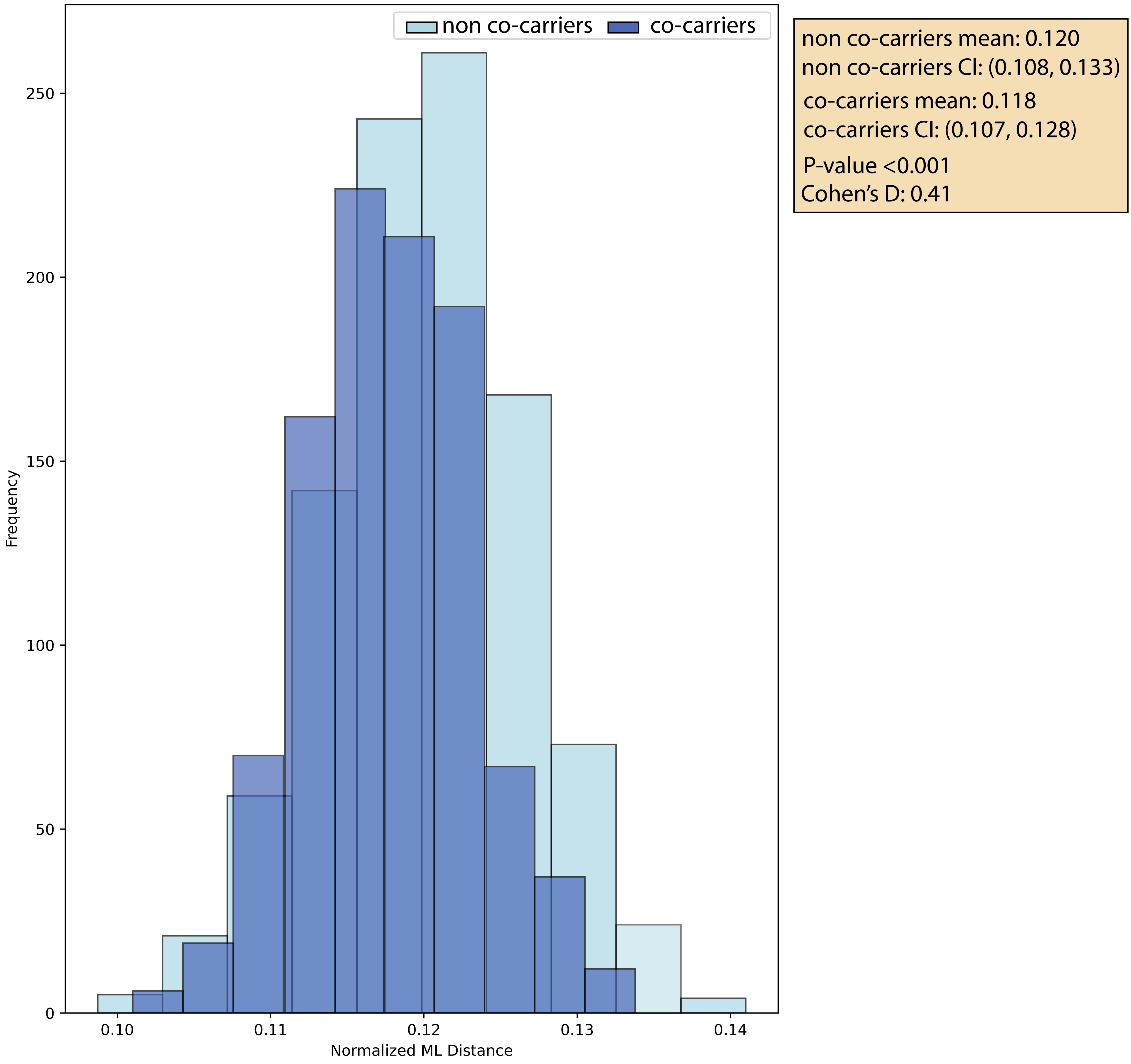
